# A Review of Surgical Education Fellowships in the United States

**DOI:** 10.1101/2025.05.12.25327475

**Authors:** Caitlin Silvestri, Maya L. Hunt, Connie Y. Gan, Michael A. Kochis, Sarah Lund, Joshua Roshal, Darian Hoagland, Ariana Naaseh, Tejas S. Sathe, Rebecca Moreci, Nicole Brooks, Riley Brian

**Affiliations:** Columbia/New York Presbyterian Hospital, Department of Surgery, New York, NY; Department of Surgery, Indiana University School of Medicine, Indianapolis, IN; Stanford University, Department of Surgery, Stanford, CA & Oregon Health and Science University, Department of Surgery, Portland, OR; Massachusetts General Hospital, Department of Surgery, Boston, MA; Department of Surgery, Mayo Clinic, Rochester, MN; University of Texas Medical Branch, Department of Surgery, Galveston, TX & Brigham and Women’s Hospital, Department of Surgery, Boston, MA; Beth Israel Deaconess Medical Center, Department of Surgery, Boston MA and Lahey Hospital & Medical Center, Simulation Center, Burlington, MA; Washington University in St. Louis School of Medicine, Department of Surgery, St. Louis, MO; Columbia University/New York Presbyterian Hospital, Department of Surgery, New York, NY; Center for Surgical Training and Research, Department of Surgery, University of Michigan Ann Arbor, MI; Department of Surgery, Cleveland Clinic, Cleveland, OH; Department of Surgery, University of California San Francisco, San Francisco CA

**Keywords:** medical education, surgical education fellowships, ACS-AEI accredited, education research

## Abstract

**Purpose:** Surgical education fellowships emerged to address the growing need for formal surgical education training. However, little is known about fellowship structure, curricula, and outcomes. This lack of transparency creates challenges for residents choosing programs, institutions developing new fellowships, and existing programs evaluating their outcomes.

**Method:** Authors identified 28 United States-based surgical education fellowships and distributed a survey to representatives (directors or fellows) from each program evaluating program structure, educational opportunities, research infrastructure, teaching experiences, and mentorship. Responses were analyzed using descriptive statistics.

**Results:** Nineteen fellowship programs (68% response rate) participated. The majority of programs offered full funding (84%). Program length varied from 1 to 2 years, most commonly with one fellow per year. Few (16%) required completion of an advanced degree during the fellowship. The majority mandated teaching (84%) and simulation (89%) responsibilities, though time commitments varied. Most programs (84%) required fellows to attend national conferences. Over half (53%) had an Education PhD advisor within the department, and 63% provided free access to statisticians. Leadership structures varied, with 74% having a single fellowship director. Among directors, 80% were surgical specialists, while all non-surgical directors held Education PhDs. Dedicated faculty time equivalents for directors varied widely.

**Conclusions:** Our findings highlight significant heterogeneity across U.S. surgical education fellowships. Transparency about fellowship structures could improve prospective fellow decision-making and guide future program development. Further research should focus on the long-term outcomes of fellowship graduates and the impact of accreditation on fellowship success.

## Introduction

Medical Education Fellowships (MEFs), first introduced in the late 1970s, were developed to address the growing need for skilled educators in medicine, particularly in Family Medicine.^1,2^ These programs provide faculty members with structured, longitudinal development in teaching methodology, curriculum design, and educational leadership.^2^ As educational science and technology advanced over the decades, re-shaping how learners are taught and assessed, MEFs offered formal training in instructional strategies, learner assessment, mentorship, and scholarly productivity. These programs also responded to the growing demand for faculty skilled in simulation, active learning, and competency-based evaluation—areas that require dedicated training beyond what is typically included in clinical education.^3,4^ Although MEFs first arose in family medicine, they have since appeared in other specialties including internal medicine, rheumatology, and emergency medicine.

In a similar vein, surgical educators have increasingly recognized the need for and importance of formal training for faculty involved in teaching, curriculum design, and academic scholarship.^5,6^ Surgeons pursuing academic careers that include education-focused roles—such as clerkship directors, residency program leadership, and simulation center faculty—require a skill set beyond clinical expertise.^7–10^ However, despite a growing interest in education, barriers such as limited time, lack of education-specific skills, inadequate mentorship, and ongoing development activities have hindered faculty development in this area.^11,12,13^

In response to these challenges, several national initiatives and courses sponsored by national organizations such as the American College of Surgeons (ACS) and Association of Surgical Education (ASE) have emerged to support surgeon-educators.^14–17^ Furthermore, Accredited Education Institutes (AEIs) were established by the ACS Division of Education in 2005 to set standards in surgical education and simulation for both faculty and trainees.^18,19^ Following the establishment of AEIs, surgical education fellowships—primarily designed for residents during their professional development time—also began to emerge in the mid-2000s, with the earliest founded in 2007.^20,21^ These were often housed within AEIs^18,19^ or other non-AEI accredited institutions,^22–24^ with the aim of providing structured training in surgical education, simulation, curriculum development, and academic scholarship.^25^

Despite the increasing number of surgical education fellowships, little is known about the structure, curricular components, graduation requirements, or fellow outcomes across these programs. The lack of information about surgical education fellowships creates significant challenges for multiple stakeholders. Residents seeking opportunities for their professional development may struggle to identify programs that align with their individual needs or to ask the right questions to understand differences between programs. For institutions aiming to establish new fellowships, the absence of global data makes it difficult to design programs that meet current needs. Existing programs, meanwhile, are hindered by a lack of data that limits their ability to evaluate their own structure and outcomes in relation to other programs. As a result, there is a critical need for a review of these fellowships to support both aspiring surgeon-educators and the institutions that train them.

Thus, the purpose of this study is to conduct a review of surgical education fellowships in the U.S. by 1) describing their breadth and diversity in structure and 2) identifying trends in advanced education opportunities, teaching, simulation, research and mentorship.

## Methods

This study was deemed exempt from full review by the Institutional Review Board at Columbia University, under Protocol #AAAV1242.

### Survey Development and Design

We developed a cross-sectional survey to evaluate the structure, educational opportunities, research infrastructure, teaching experiences, and mentorship within accredited and non-accredited U.S. surgical education fellowships. We followed established best practices for survey development to create a survey based on a review of existing fellowship structures to ensure accuracy and relevancy of the content.^26^ We piloted the survey with fellowship directors and members of the Collaboration of Surgical Education Fellows (CoSEF) network, which includes general surgery residents with an interest in education, as well as former and current surgical education fellows. Based on pilot testing, the survey was iteratively revised using feedback from stakeholders. The final survey consisted of 72 items and included a mix of open- and close-ended questions. The complete survey can be found in the Supplementary Information (Supplementary A).

### Survey Content

Prior to starting the survey, we obtained participant demographics including position (current or former fellow, fellowship director) and number of years since program graduation, if applicable.

The survey consisted of five major domains including 1) program structure, 2) advanced educational opportunities, 3) teaching and simulation experience, 4) research infrastructure and 5) mentorship and leadership (Figure 1).

**Figure 1.**
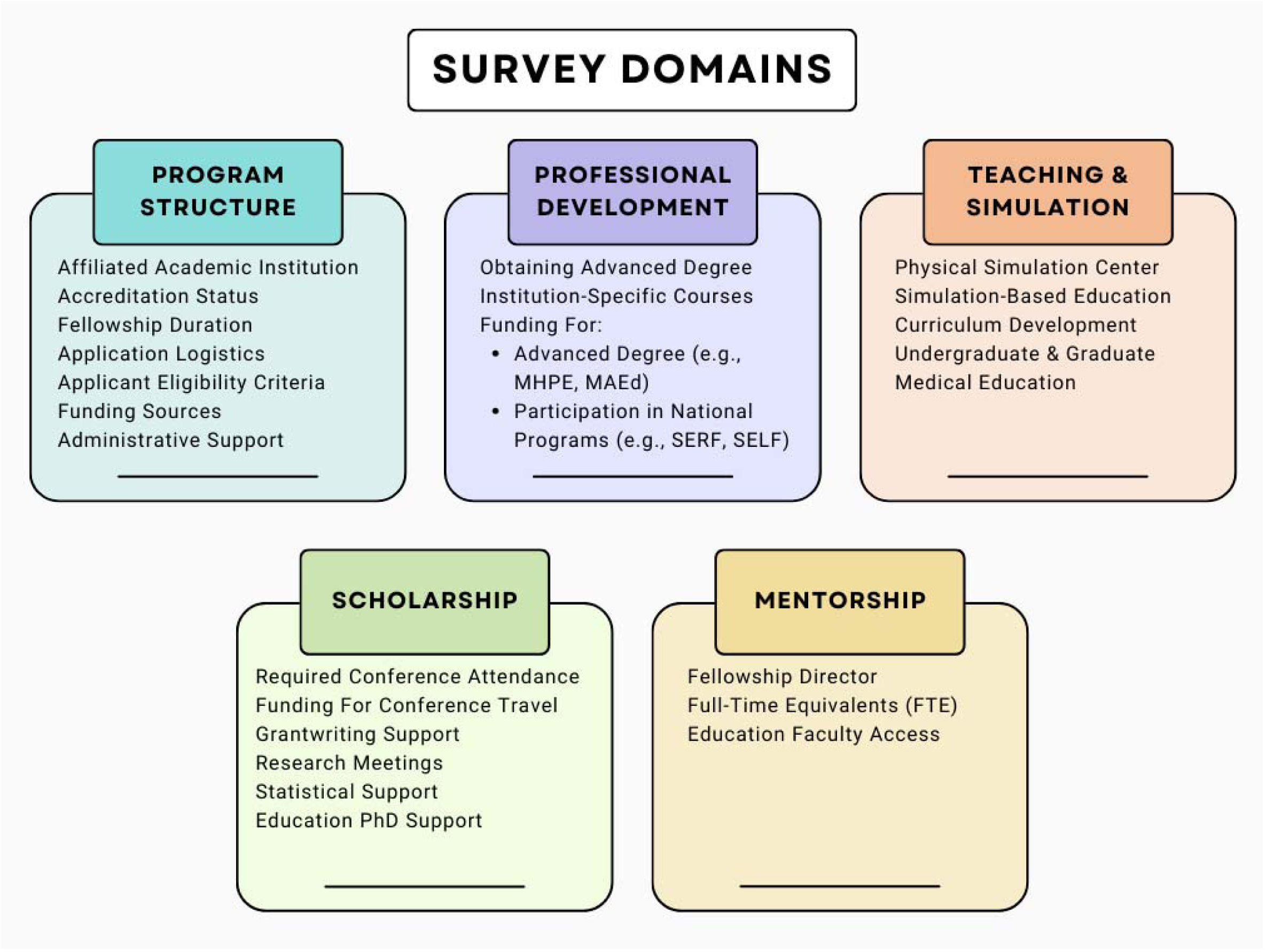
Five survey domains and affiliated subcategories.

### Participants

We identified surgical education fellowships in the U.S. by querying the ACS-accredited fellowship list,^25^ CoSEF,^27^ additional web searches, and the ACS-AEI Surgical Simulation Summit Conference registration. Among non-ACS-AE accredited fellowships, we excluded those that (1) were accredited by other organizations (e.g., the Society for Simulation in Healthcare), (2) focused exclusively on simulation without broader educational components, or (3) did not include a primary focus on surgical education or simulation.

We then identified representatives including current fellowship directors, as well as current and any past fellows for survey distribution. We identified possible participants via publicly available websites and in-person recruitment at the ACS-AEI Simulation Summit Conference. After identifying all possible representatives of a program, we then sent a single invitation email to all representatives identified to participate. While only one representative was required to complete the survey, we encouraged them to collaborate with others at their program to ensure the responses were complete and accurate.

### Survey Administration and Distribution

We developed the survey and distributed it using Tally, an online survey platform (Tally BV, Ghent, Belgium), with responses collected from April 2024 to December 2024. We sent two email reminders to encourage participation. Participation was voluntary, and no incentives were offered for survey completion.

### Data Analysis

We utilized Microsoft Excel (Version 16.78, Microsoft Corporation, Redmond, WA) for descriptive statistical analysis Categorical variables were reported as frequencies and percentages; continuous variables were summarized with means, medians, ranges, with standard deviation and interquartile ranges as appropriate.

## Results

### Demographics and Response Rates

We identified a total of 28 fellowship programs, with 20 accredited and 8 non-accredited fellowships. Representatives of 19 fellowships completed the survey, (overall response rate 68%) including 15 accredited programs (75% response rate) and 4 non-accredited programs (50% response rate). Respondent demographics included 17 (89%) fellows and 2 (11%) fellowship directors. Among fellows who responded, 14 (74%) were current fellows, 2 (11%) graduated one year prior, and 1 (5%) graduated over two years ago.

### Program Overview and Logistics

The programs analyzed were established between 2007 and 2023, with the largest proportion established in 2018 (*n*=5) (Figure 2). Most programs were located in the Midwest (*n*=8, 42%). Fellowship program lengths vary, with an approximately equal distribution among one year, flexible 1 or 2 year, and two year programs. Most progra 84%) accept applications annually, while the remaining programs had varying application cycles (Table 1).

**Figure 2.**
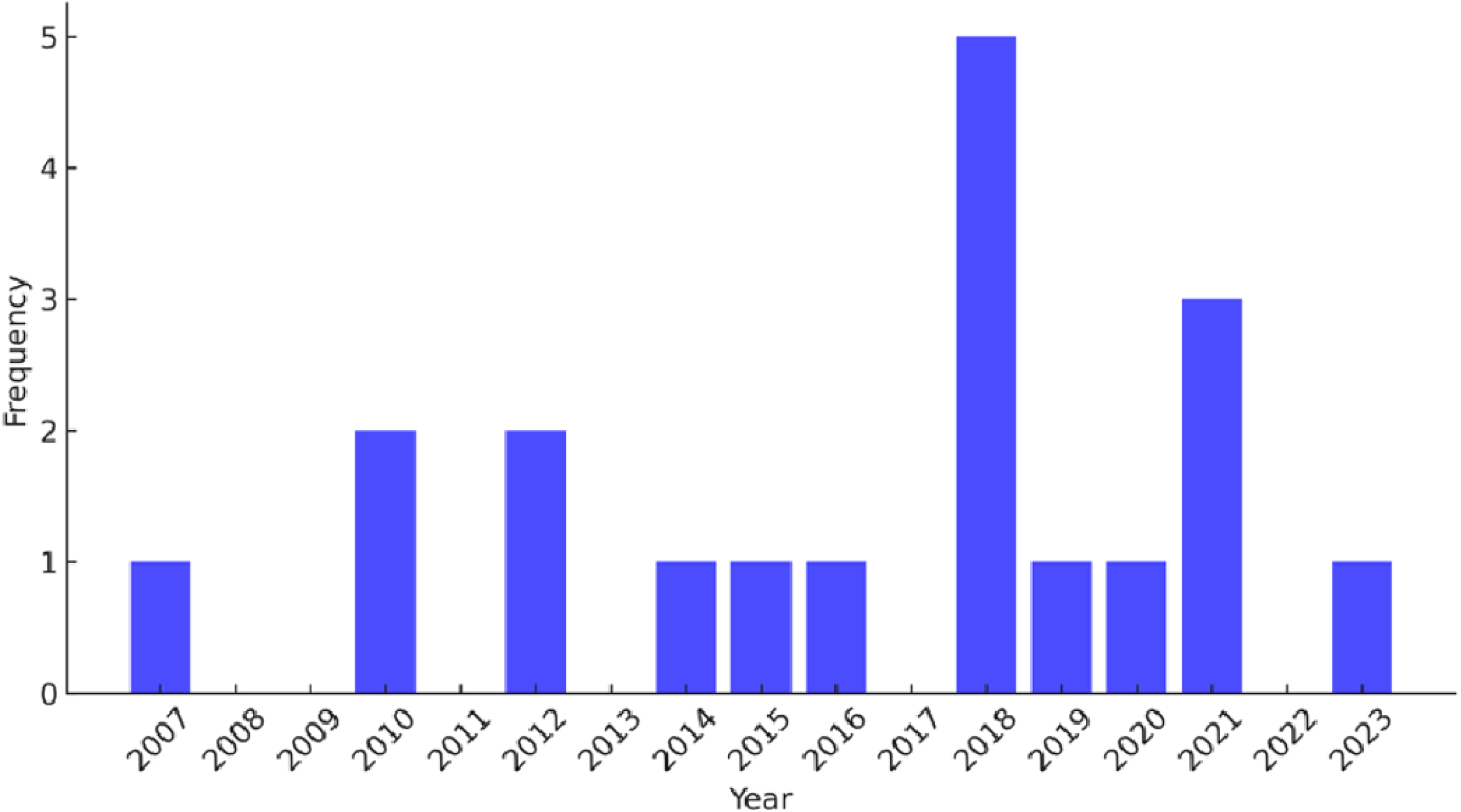
Number of fellowship programs established each year.

**Table 1.** Overview of fellowship program characteristics and logistics.

| **Fellowship Program Characteristic** | **Frequency (%)** |
| --- | --- |
| *Program Location (n=19)* |  |
| Midwest | 8 (42%) |
| Northeast | 5 (26%) |
| West | 3 (16%) |
| Southwest | 2 (11%) |
| Southeast | 1 (5%) |
| *Fellowship Length (n=19)* |  |
| 1 year | 6 (31.5%) |
| 1 or 2 years | 7 (37%) |
| 2 years | 6 (31.5%) |
| *Application Cycle (n=19)* |  |
| Every year | 16 (84%) |
| Every 1 to 2 years | 1 (5%) |
| Every 2 years | 2 (11%) |
| *Positions reserved for internal candidates (n=19)* |  |
| Yes | 8 (42%) |
| No | 8 (42%) |
| Unsure | 3 (16%) |
| *Percentage of internal candidates between 2018-2023 (n=18)* |  |
| 100% | 5 (28%) |
| 75-99% | 3 (17%) |
| 50-74% | 4 (22%) |
| 25-49% | 1 (5%) |
| 1-25% | 2 (11%) |
| 0% | 3 (17%) |
| *Types of applicants accepted (n=19)* |  |
| General surgery or surgical subspecialties only | 14 (74%) |
| Surgical and non-surgical | 5 (26%) |
| *International applicants (n=19)* |  |
| Yes | 8 (42%) |
| No | 10 (53%) |
| Unsure | 1 (5%) |
| *Fully remote position available (n=19)* |  |
| Yes | 4 (21%) |
| No | 15 (79%) |
| *Fully funded position (n=19)* |  |
| Yes | 16 (84%) |
| No | 3 (16%) |

From 2018 to 2023 (5 academic years), programs reported a median of 5 fellows (IQR 3, 7.75), with a range of 0–16 fellows per program. The single program that reported 0 fellows was newly established in 2023 and had not yet begun recruiting. Programs are often associated with a hospital or university, and half of programs (*n*=8, 42%) report reserving fellowship positions for internal candidates. Additionally, among programs with fellows during this period, 5 (28%) had exclusively accepted internal fellows and 7 (39%) had internal candidates comprise more than half of their fellow cohort. In terms of applicant specialty, 7 programs (37%) limit eligibility exclusively to general surgery applicants, while another 7 (37%) are open to all surgical subspecialties, resulting in 74% of programs restricting participation to surgical trainees. Eight programs (42%) consider international applicants. Lastly, 4 programs (21%) offer fully remote positions, allowing fellows to participate in the fellowship without being physically located in the same state as the fellowship institution (Table 1).

The majority of programs provide full salary funding for the fellows (*n*=16, 84%) (Table 1). Among fully funded programs, the median salary was $67,000. Most salaries were reported to be based on the corresponding PGY salary at that institution, making these figures representative of salaries during the 2023–2024 academic year. Funding for programs and fellows came from a variety of sources, including the department of surgery, medical school, simulation center, industry, and cost-sharing with the external fellows’ home institution. Many programs reported receiving funding from multiple sources.

### Professional Development

Prior to starting the fellowship, 17 programs (90%) provide their fellows with written goals and objectives. Pursuit of an advanced degree, usually Master of Health Professions Education or Masters in Education, is required in 3 program (16%) and highly encouraged in another 5 (26%). Notably, all programs that mandate an advanced degree provide full financial support for tuition (*n*=3, 100%). Among programs that highly encourage an advanced degree, 4 (80%) offer full tuition coverage, while 1 (20%) provides partial financial support. In contrast, programs that do not require or highly encourage an advanced degree also do not offer funding for fellows who choose to pursue one independently.

While programs do not typically require an advanced degree, the majority have had prior fellows participate in organizational scholarship programs such as the Association of Surgical Education (ASE) Surgical Education Research Fellowship (SERF) or Leadership Fellowship (SELF), with 12 (63%) and 4 (21%) having at least one fellow participate in each program respectively. Over half of programs (*n*=10, 53%) reported providing full funding for fellows to attend SERF or SELF.

### Teaching and Simulation

Among fellowships, a substantial proportion incorporate simulation and teaching responsibilities within undergraduate (UME) and graduate medical education (GME) spaces. The majority of programs (*n*=18, 95%) provide access to a simulation center, and 17 programs (89%) require fellows to participate in the administrative coordination and direct instruction of simulation sessions. Regarding teaching, 16 programs (84%) have teaching obligations that vary widely in audience and scope. Among these programs, UME teaching commitments ranged from 4 to 30 hours per month (median 10.0 hours, [IQR 4,10]), while GME teaching commitments ranged from 0 to 30 hours per month (median 10.0 hours, [IQR 4,12]). Fifteen (79%) programs require fellows to participate in curriculum development. The majority of programs offer opportunities for fellows to develop expertise in teaching, curriculum design, and education research methodology through institutional courses (*n*=14, 74%). Lastly, administrative support varies, with 15 programs (79%) employing a full- or part-time administrator specifically for the fellowship.

### Research

Surgical education fellows frequently engage in networking, collaboration, and research as part of their training. A majority of programs (*n*=16, 84%) require fellows to attend national conferences, with the ACS-AEI Simulation Summit being the most common (*n*=12), followed by ASE and Association of Program Directors in Surgery (APDS) meetings (*n*=8) (Table 2).

**Table 2.** Fellowship program research requirements and institutional support.

| **Fellowship Program Characteristic** | **Frequency (%)** |
| --- | --- |
| *Require attendance at national conference (n=19)*<br>Yes<br>No | 16 (84%)<br>3 (16%) |
| *Require fellows submit a research grant proposal for external funding (n=19)*<br>Yes<br>No | 3 (16%)<br>16 (84%) |
| *Financial support for required conferences (n=16)*<br>Full reimbursement<br>Partial reimbursement<br>No reimbursement | 13 (81%)<br>2 (13%)<br>1 (6%) |
| *Financial support for conferences at which fellow is presenting (n=19)*<br>Full reimbursement<br>Partial reimbursement<br>No reimbursement | 16 (84%)<br>2 (11%)<br>1 (5%) |
| *Financial support for optional conferences (n=19)*<br>Full reimbursement<br>Partial reimbursement<br>No reimbursement<br>Unsure | 4 (21%)<br>3 (16%)<br>11 (58%)<br>1 (5%) |
| *Access to an Education PhD within department of surgery (n=19)*<br>Yes<br>No | 10 (53%)<br>9 (47%) |
| *Access to an Education PhD within the institution (n=9)*<br>Yes<br>No | 4 (44%)<br>5 (56%) |
| *Access to a statistician at no cost (n=19)*<br>Yes<br>No<br>Unsure | 12 (63%)<br>6 (32%)<br>1 (5%) |

Financial support for conference attendance varies across programs. When fellows attend a required conference, 13 programs (81%) provide full reimbursement, while 2 (13%) offer partial reimbursement. When fellows present at a conference, 16 programs (84%) fully reimburse expenses. However, funding is more limited for optional conferences where fellows are not presenting, with only 4 (21%) providing full reimbursement and another 3 (16%) offering partial support (Table 2).

Regarding grant application experience, 3 programs (16%) mandate that fellows submit a research grant proposal for external research funding during their fellowship (Table 2). Among programs without this requirement, 11 (69%) have had fellows voluntarily apply for research grants. For programs that have had fellows submit grant applications, all (*n*=14, 100%) have successfully received funding at least once.

Institutional support for research was accessed by asking about access to doctoral-trained education scientists and statistical resources. Over half of programs (*n*=10, 53%) have an Education PhD mentor associated with the fellowship Of programs that do not, 4 (44%) have one accessible within the institution if needed. Regarding statistical support, 12 programs (63%) offer fellows access to statisticians at no cost (Table 2).

### Fellowship Directors

Fellowship leadership structure varies across programs with the majority (*n*=14, 74%) having a single fellowship director and 5 (26%) having greater than one. Among all fellowship directors, 20 (80%) were from surgical specialties, while non-surgical directors were exclusively (*n*=4, 100%) Education PhDs.

Fellowship director time allocation to the program varies considerably. Among all fellowship directors with full-time equivalent (FTE) data available *(n*=21), FTEs ranged from 0-1, with an average of 0.44 (SD 0.44) and median of 0.2 (IQR 0,1). When stratified by specialty, surgeon fellowship directors (*n*=18) had a similar FTE range of 0-1 and average of 0.42 (SD 0.42) and median of 0.2 (IQR 0, 1). Education PhD fellowship directors (*n*=3) had a similar FTE range of 0-1 with a slightly higher mean of 0.5 (SD 0.71) and median of 0.5 (IQR 0.25, 0.75), but greater variability compared to their surgeon counterparts.

Mentorship within the fellowship was a strong component of the program structure. In the majority of programs (*n*=17, 89%), the fellowship director serves as the primary mentor for fellows. Fifteen programs (79%) provide additional mentors to support fellows’ professional and academic development.

Additional findings from the survey have been collated in Supplementary Table B.

## Discussion

To the best of our knowledge, this study provides the most comprehensive analysis of surgical education fellowship programs across the United States. While there are several commonalities among programs—including potential advanced education, hands-on teaching and simulation, and research—our findings highlight substantial heterogeneity in program structure, resources, and opportunities for fellows. This variation underscores the need for a clear framework to guide prospective applicants in selecting programs that align with their career goals and individualized needs.

In addition to guiding applicants, our report holds important implications for various stakeholders in academic surgery. For educators developing new fellowships, recognizing the diversity of existing programs can provide a curricular foundation and inspire innovative approaches to surgical education. Drawing on these findings can help inform the creation of intentional, mission-driven curricula that align with the unique goals of each institution. Similarly, for established programs, our results offer an opportunity for reflection and benchmarking—to ensure continued alignment of their missions with evolving priorities and the broader landscape of surgical education.

### Growth and Institutional Trends

The number of surgical education fellowships has nearly doubled in the last decade, with a notable surge in new programs since 2018. This growth likely reflects increasing interest among trainees in formal surgical education training, expansion of simulation technology in surgery, as well as a broader recognition of surgical education as an academic career pathway.^5,6^ These fellowships are geographically diverse, with varying program lengths, application cycles, and eligibility criteria. A key finding is the tendency for programs to prioritize internal applicants, denoted by reports of reserving spots for internal applications and a significant proportion of fellows coming from within their home institution. This may be for a variety of reasons other than explicit preferential selection of internal candidates, such as financial constraints or a lack of external visibility of these fellowships. It may also be the result of self-selection bias in which medical students with a pre-defined interest in education specifically seek out residency programs with education fellowships, thus creating a residency cohort naturally predisposed to pursue these programs during their professional development time. A centralized location for information on both accredited and non-accredited programs with website links and program contacts may improve awareness, access, and equity for trainees nationally.^28–30^ Individual programs can also consider targeted outreach efforts to trainees outside their institutions, akin to recruitment strategies employed by clinical subspecialty fellowships.^31,32^

### Professional Development and Advanced Education

A minority of programs *require* fellows to pursue an advanced degree during their fellowship time, while additional programs strongly encourage it and often provide full financial support for fellows. Because a Master’s in Education typically costs around $44,640,^33^ programs that require or strongly encourage fellows to pursue an advanced degree— and offer full financial support—represent a substantial professional and financial investment in their fellows. A Master of Education (MEd) or Master of Health Professions Education (MHPE) degree exposes fellows to formal training in curriculum design and evaluation, research methods, and educational leadership.^34,35^ The proposed values of these programs are improved pedagogical knowledge, mentorship and development of networks, increased educational scholarship, and broader career opportunities, though the value will inherently be individualized.^36,37^ The pursuit of an advanced education degree specifically during professional development time is associated with increased research productivity and impact factor within academic surgery.^38^ While such degrees are not necessarily required, they may better prepare fellows for careers in academic surgery such as medical school clerkship director, residency or fellowship program director, and education scholars.^39^

Aside from advanced degrees, many programs supported trainee participation in ASE’s SERF and SELF educational programs. Such programs may provide the foundational knowledge and skills, mentorship, and networking opportunities on a smaller scale that may integrate more easily into the specific structure of each fellowship. Although tuition costs for these programs are lower than formal degrees,^40^ they still represent a meaningful financial investment by the program in the fellow’s development.

If prospective fellows are interested in an advanced degree during their professional development time, the degree of financial support provided by potential fellowship programs is essential. In turn, programs should critically reflect on their own goals, resources, and limitations to define the strongest version of their fellowship. This includes evaluating whether requiring or supporting advanced degree completion aligns with their mission.

### Teaching and Simulation-Based Education

Teaching and simulation-based education are central components of surgical education fellowships, yet the degree of fellows’ hands-on involvement–and the support provided to them–varies widely. Nearly all programs require fellows to coordinate and lead simulation sessions, and many also mandate substantial teaching commitments at both the UME and GME levels. Programs with heavier teaching loads may provide stronger preparation for future roles in academic surgery, particularly for trainees seeking careers as clinician-educators.

However, the time demands, quality, and level of support for these teaching responsibilities can vary significantly across programs. The extent of formal training in education is variable with inconsistent expectations and availability across programs regarding structured coursework through institutional offerings, degree programs, or national courses and initiatives.^41–43^ Given the central role fellows play in education delivery, it is important to consider whether programs are providing the necessary tools and mentorship for trainees to grow into these responsibilities. Fellows are still developing as educators, and while taking on significant teaching roles can be valuable, it should be accompanied by intentional training and graduated support.^44^ Ensuring that teaching responsibilities are aligned with appropriate preparation may enhance both fellow development and the quality of education they provide.

### Research and Scholarly Productivity

Scholarship expectations and support also differ among fellowships. While few programs mandate grant submissions, many fellows voluntarily pursue research funding with a high success rate. However, variable conference attendance requirements and financial support for conference travel may hinder a fellow’s ability to disseminate their work and engage or network with the broader education research community. Notably, attendance at the most frequently reported conference—the ACS-AEI Simulation Summit—is a requirement for fellowship accreditation, which may skew the data toward the experiences of fellows in accredited programs. Programs that provide robust research infrastructure, expectations, and resources may better position fellows for success in conducting high quality research within academic surgical environments. Prior studies have shown that residencies who have research programs that incorporate research requirements (e.g., conference attendance, manuscript submissions) and have readily accessible administrative support (e.g., grant application, statistical support) increase resident productivity.^45^ Not only the expectations for research, but also the support that is provided to fellows, should be considered based on an applicant’s individual needs and goals as well as the objectives of the program.

### Mentorship and Leadership Structure

Mentorship plays a pivotal role in the fellowship experience. Nearly all programs designate the fellowship director as the primary mentor, with many offering additional faculty mentorship. While most programs have a single director, others employ a co-directorship model, often incorporating non-surgical faculty such as Education PhDs. Time commitment to the fellowship also varies, as demonstrated by our finding that surgeon fellowship directors receive less FTE compared to their Education PhD counterparts. This discrepancy may impact the depth of involvement and mentorship, program development, and fellow support, as studies have shown a correlation between providing FTE to faculty for educational roles and increased participation in education activities and mentoring.^46^

### Limitations and Future Directions

Our study has several limitations. First, while survey respondents were instructed to complete the survey collaboratively with the fellow, fellowship director, and fellowship coordinator when applicable, we were unable to reliably determine whether responses were cross-checked among multiple sources. As a result, the data may primarily reflect self-reported information, predominantly from fellows rather than program directors, which may introduce inaccuracies. Additionally, certain survey questions allowed respondents to select “unsure” as an answer choice, which limits the overall sample size for those questions. Third, the small number of surgical education fellowships nationwide prohibited meaningful statistical comparisons between accredited and non-accredited programs in aggregate. Finally, our cross-sectional survey did not account for how fellowship offerings may change over time. Given that the study was conducted in 2024, the responses may not fully reflect the current structure, funding, or opportunities available in these programs at the time of publication.

Future research should collect complete data on all surgical education fellowships, with particular attention to non-accredited programs to determine whether accreditation influences fellowship structure and support or even fellowship outcomes. Establishing a comprehensive, centralized database that includes both accredited and non-accredited fellowships—along with key details such as program websites and contact information—may help standardize access to information, increase program transparency, and support more equitable opportunities for prospective applicants. Research on MEFs has explored their long-term impact on graduates’ careers, including leadership attainment, research productivity, and academic promotion.^47^ These studies provide a valuable model for future research on surgical education fellowships, which remain under-investigated in this regard. Similar to the MEF literature, tracking surgical education graduates’ career trajectories would offer important insight into the fellowship’s influence on academic development and educational leadership. Another key area of prior MEF research has been the perceived value of fellowship training among both education leaders and trainees.^8,48^ Future studies should explore how surgical education fellows and program directors assess the value of fellowship training, and how their priorities shape the structure and goals of these programs. For example, a well-executed study could help clarify whether the current heterogeneity across surgical education reflects the diverse aspirations of education-focused surgeons or signals a need for more centralized and standardized training.

## Conclusion

Surgical education fellowships play a vital role in preparing the next generation of surgical educators. This study’s findings reveal significant variability in program structures, professional development opportunities, teaching requirements, research expectations, and mentorship. While this heterogeneity can enable fellows to select training opportunities that are uniquely suited to their individual career goals, greater standardization in certain core areas— such as formalized instruction in pedagogy, simulation education, and scholarship—may enhance the overall fellowship experience. As surgical education continues to evolve as an academic discipline, further research is needed to determine the long-term career trajectories of fellowship graduates, the impact of program accreditation, and the role of various support structures in influencing fellowship outcomes.

## Data Availability

All data produced in the present study are available upon reasonable request to the authors

## Disclosures

### Acknowledgments

None.

### Disclaimers

None.

### Previous presentations

Presented at Society of Asian Academic Surgeons (SAAS) Conference on September 4th, 2024 in New Orleans, LA.

### Data

The data analyzed in this study were collected directly by the authors through an original survey distributed to representatives of surgical education fellowship programs. No external or previously published datasets were used.

