## Supplementary figures and images for "A Review of Surgical Education Fellowships in the United States"

### Supplemental A

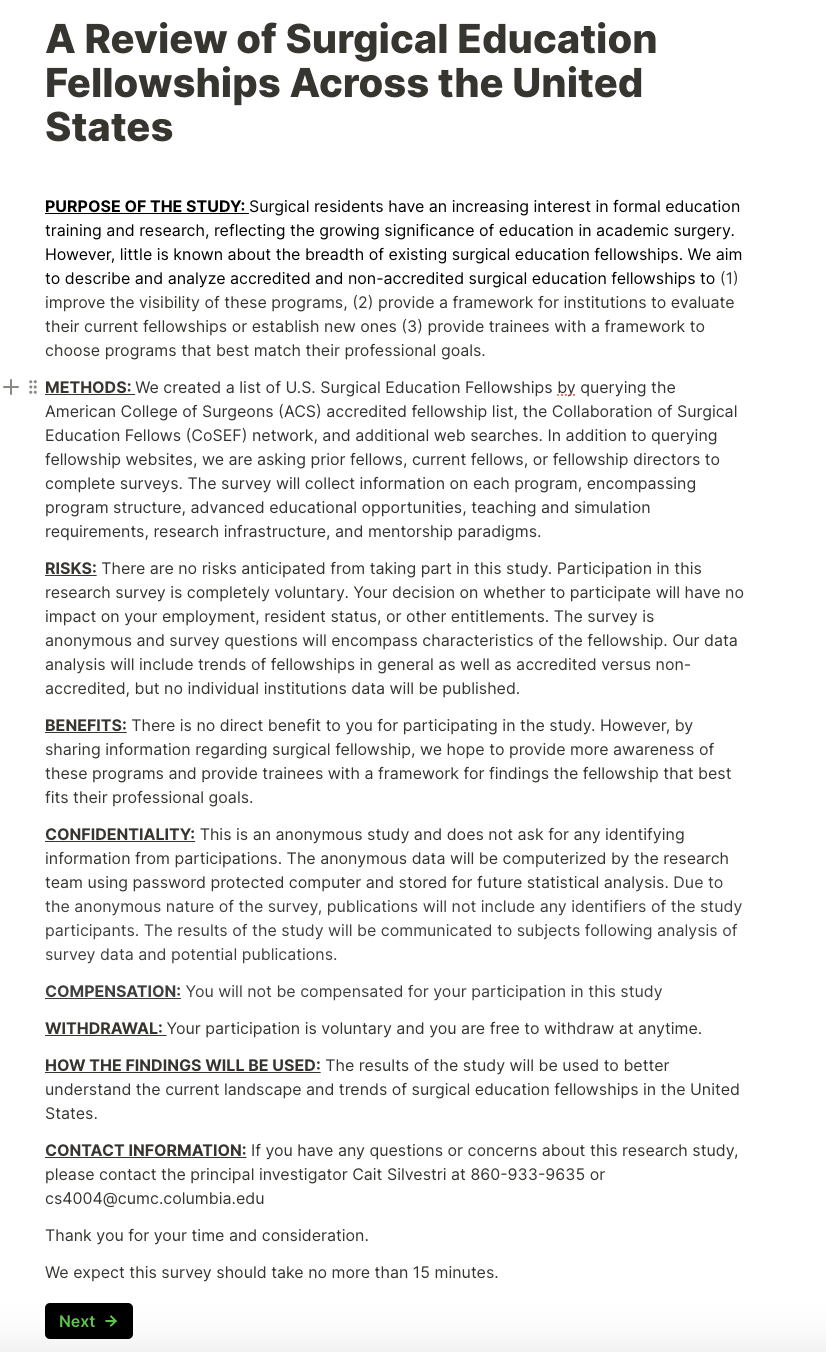

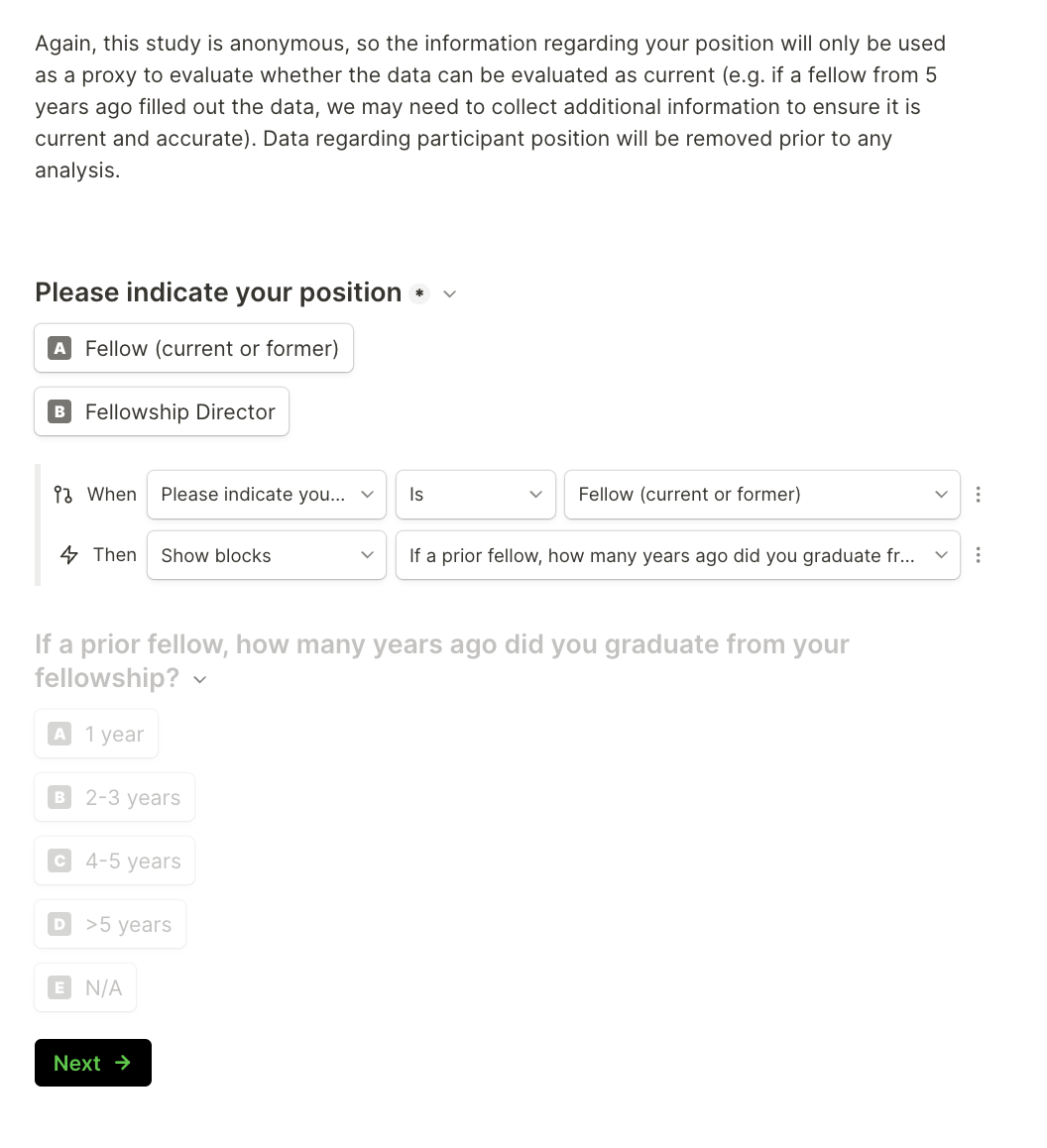

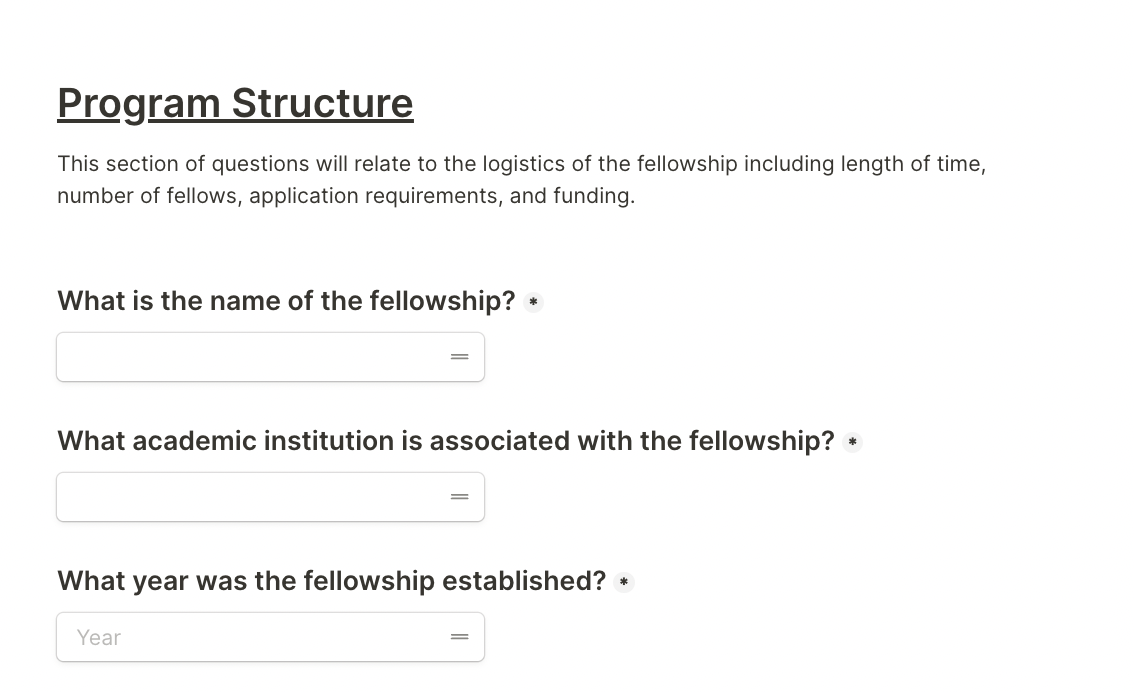

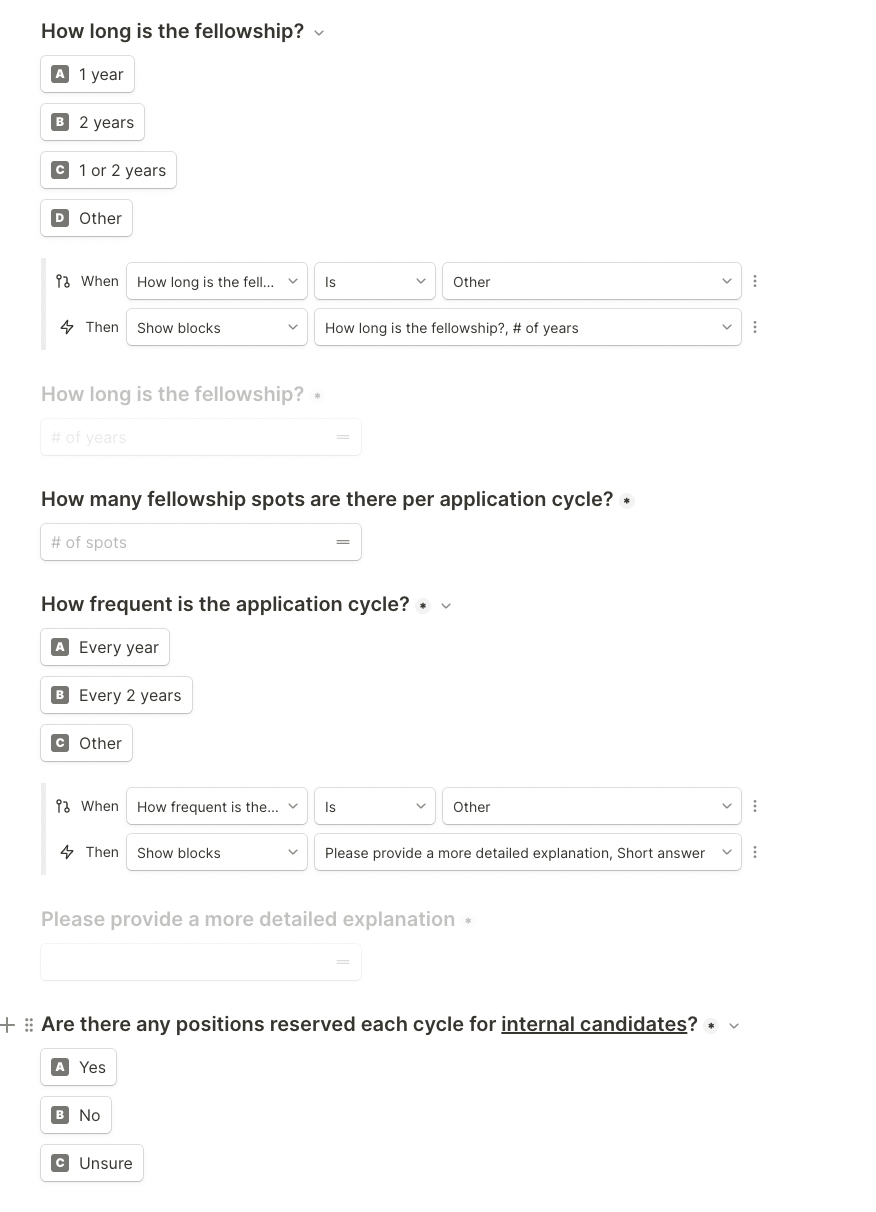

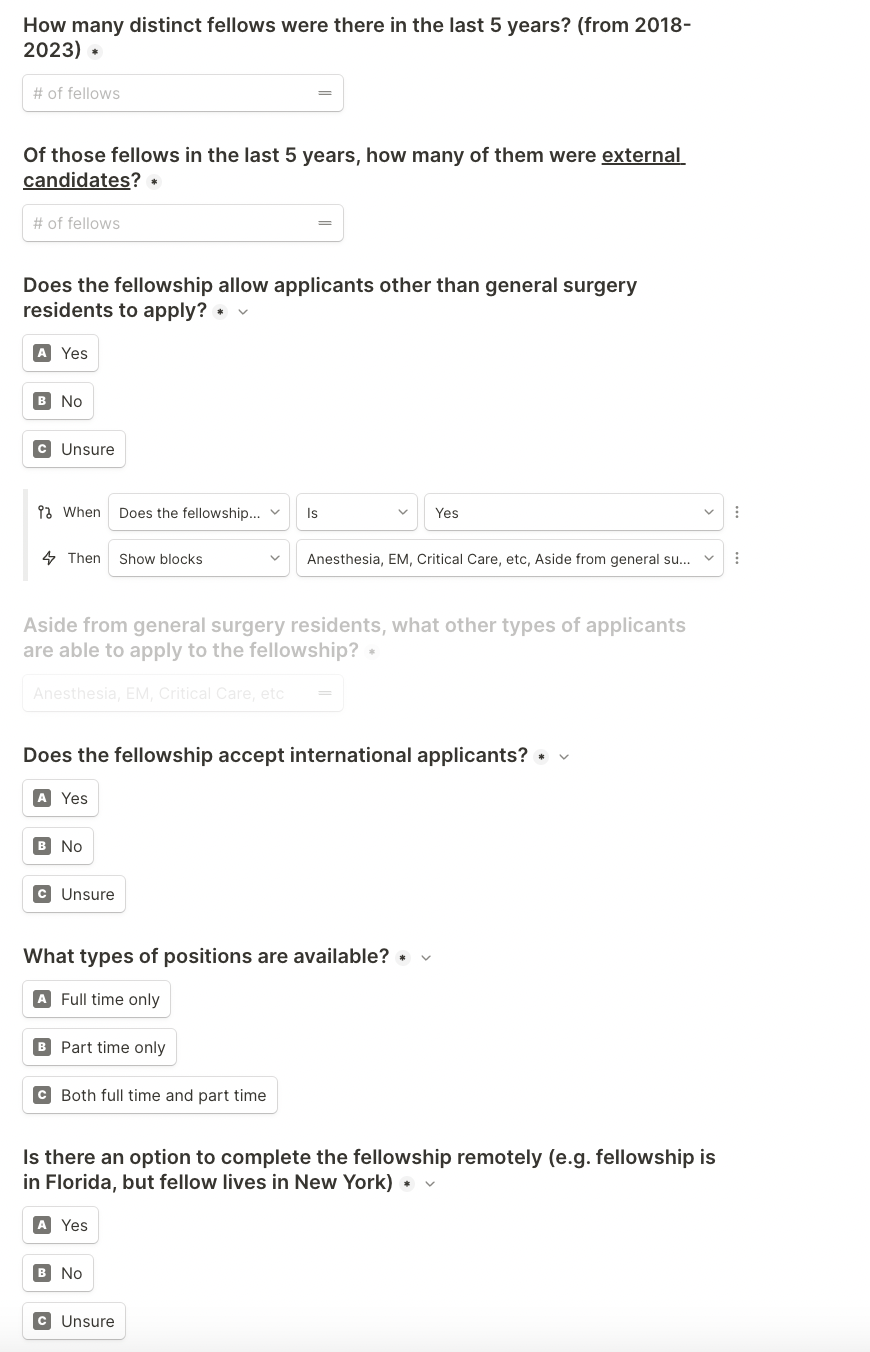

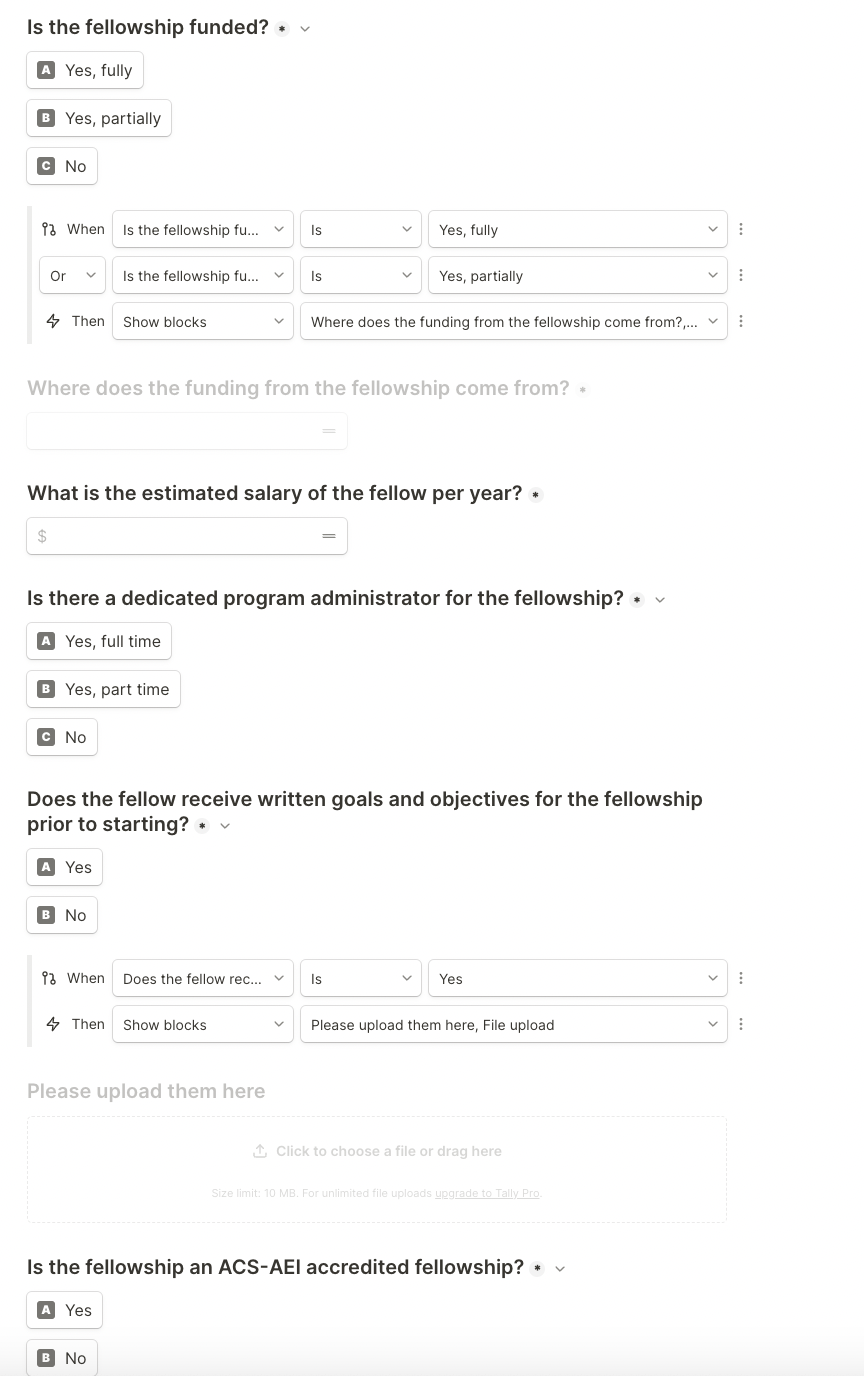

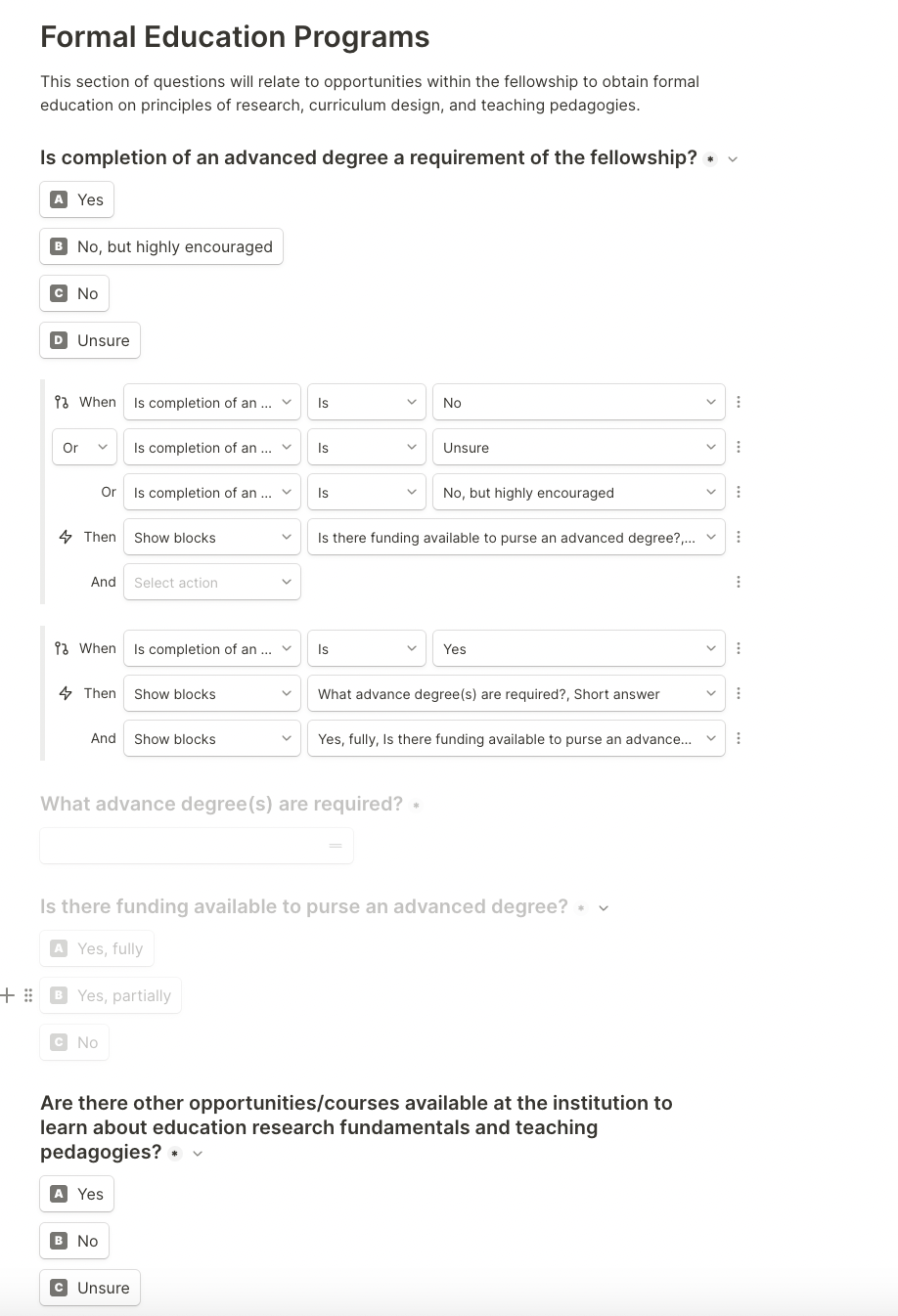

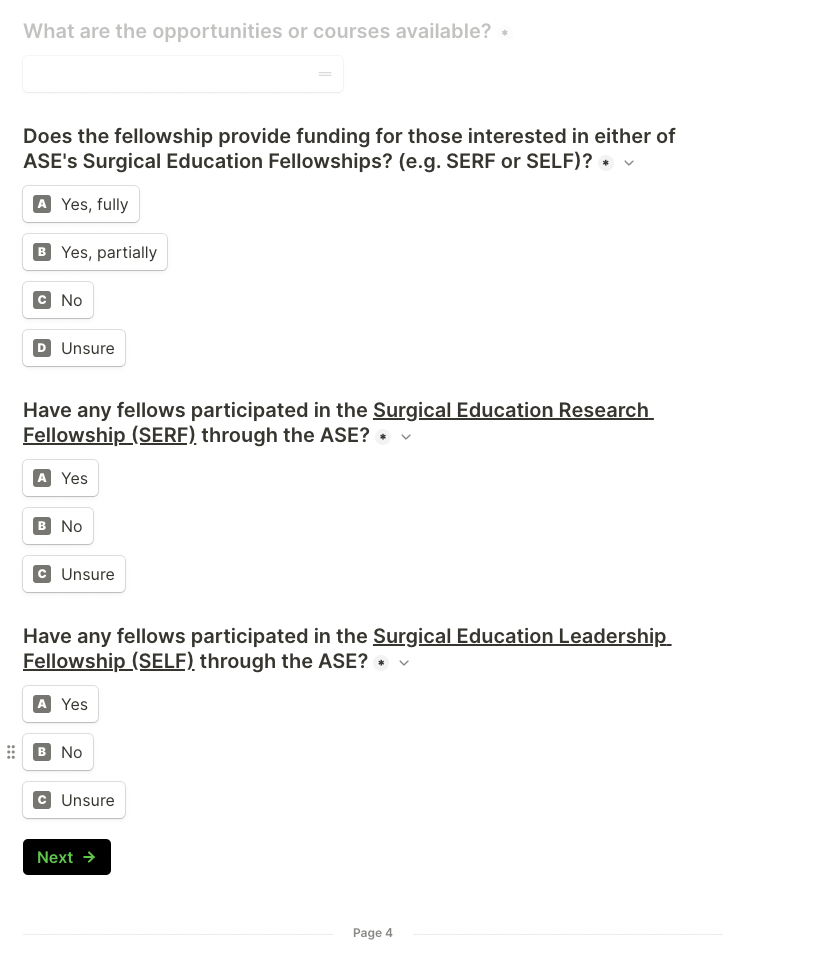

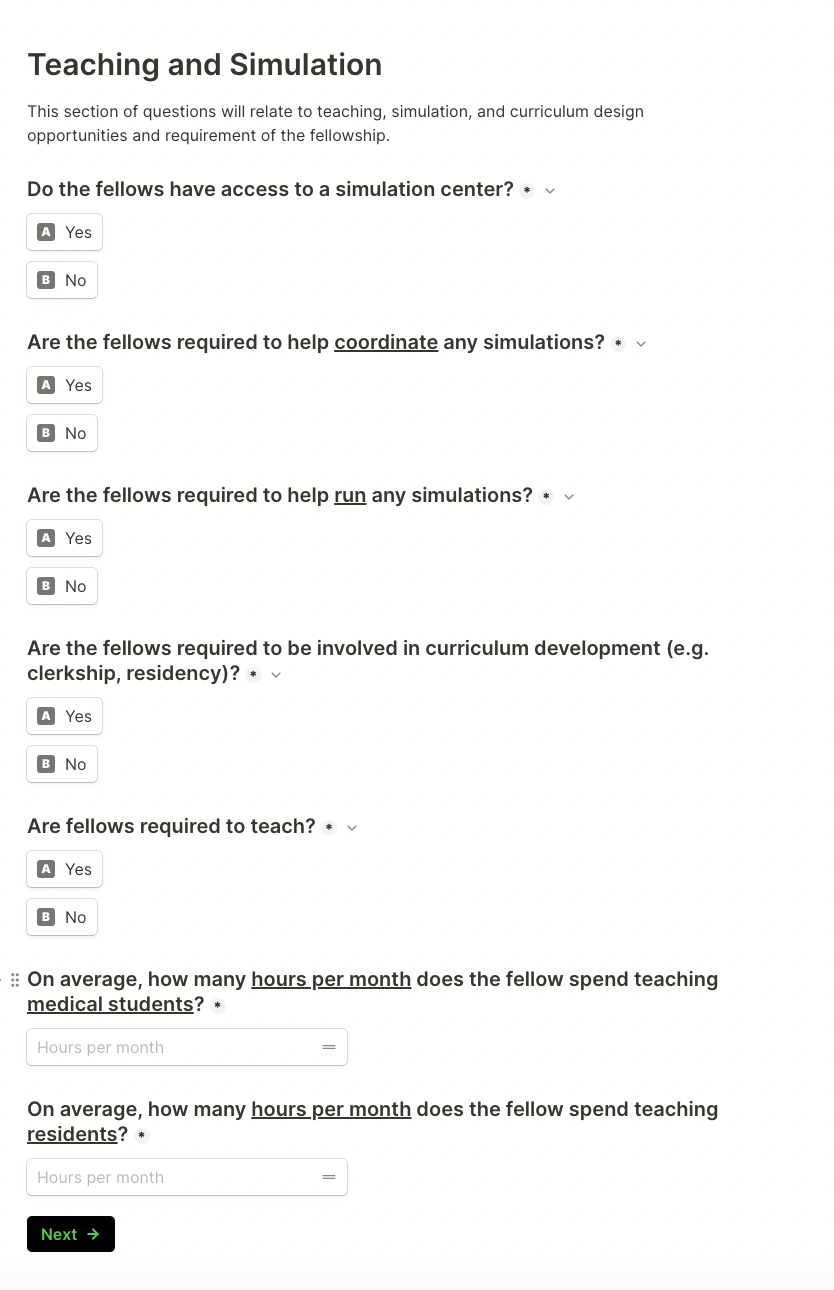

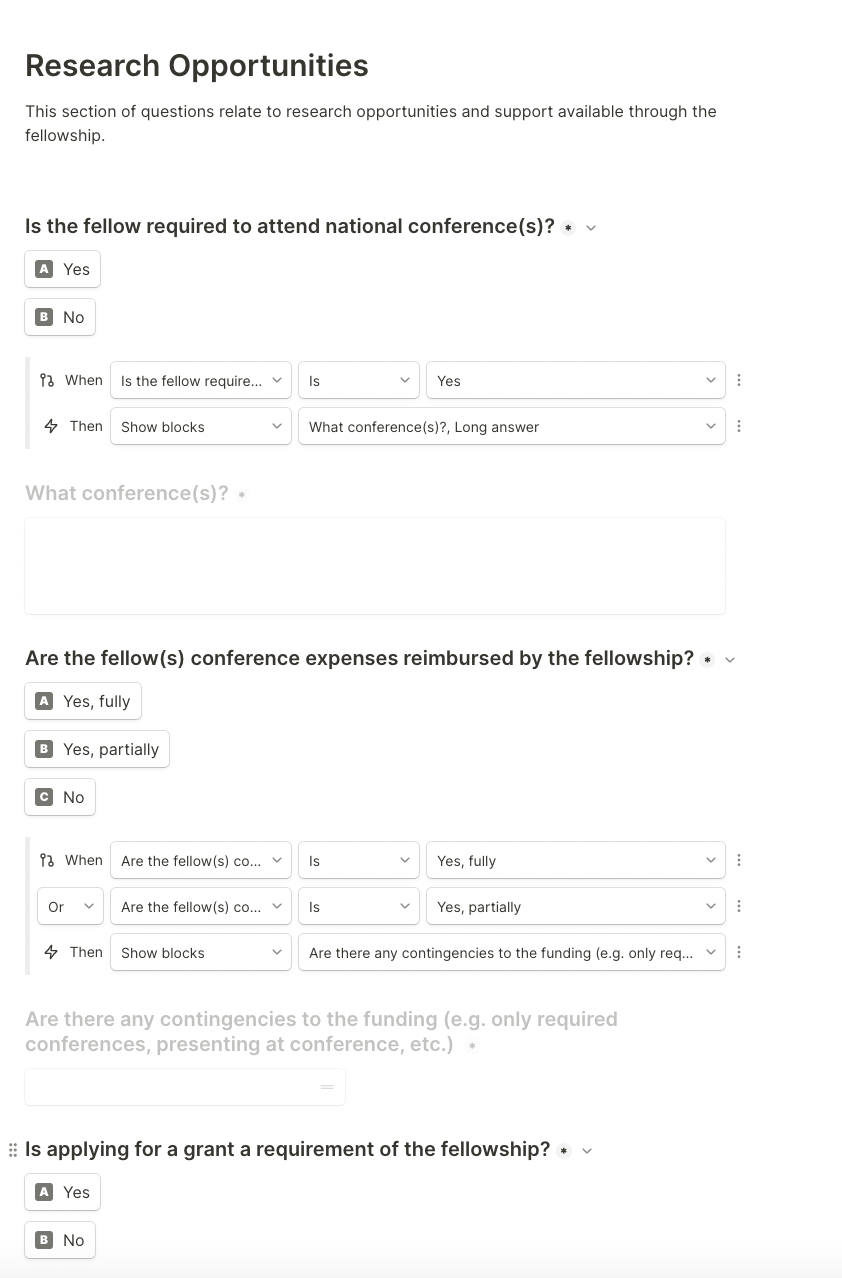

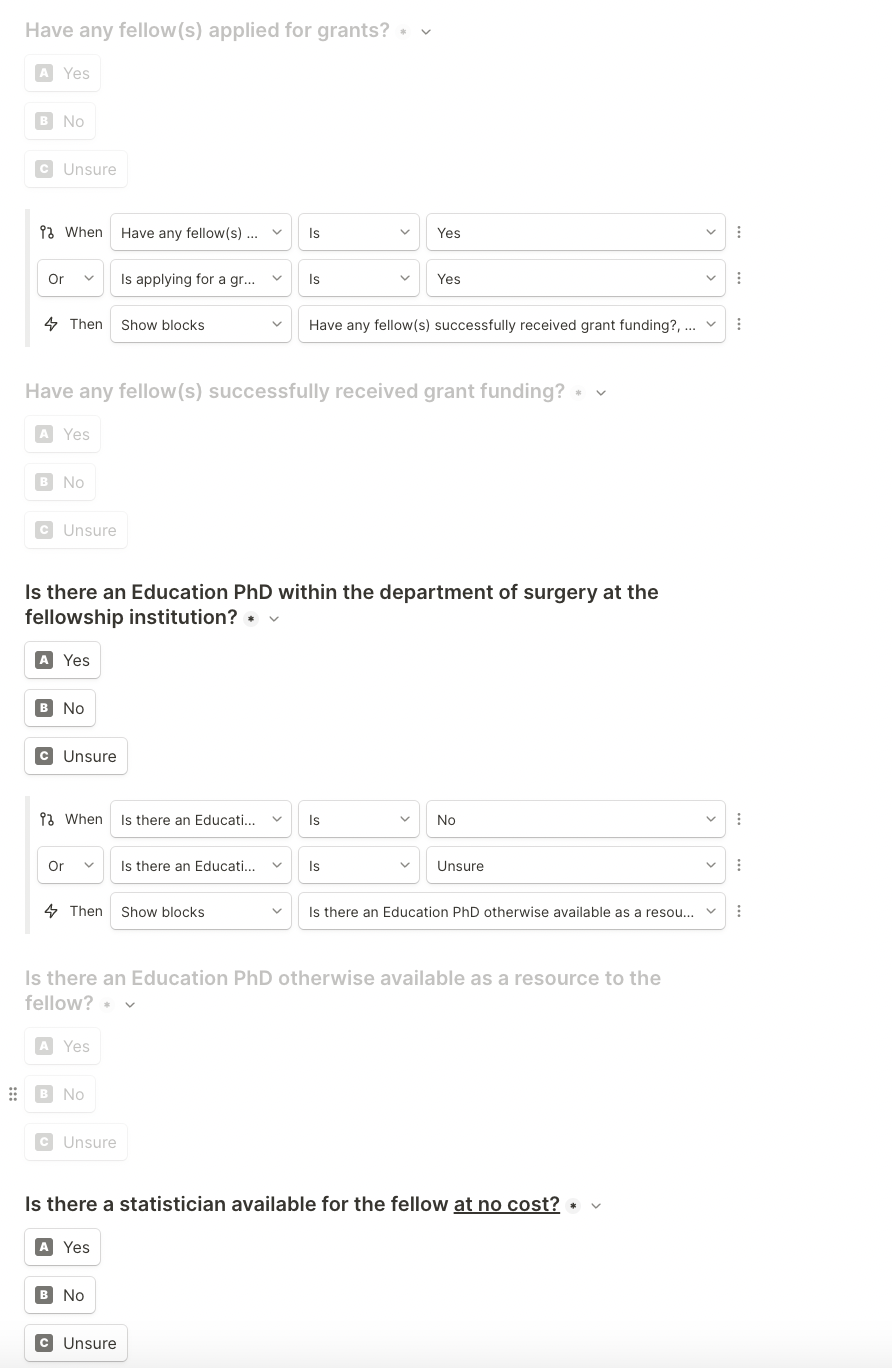

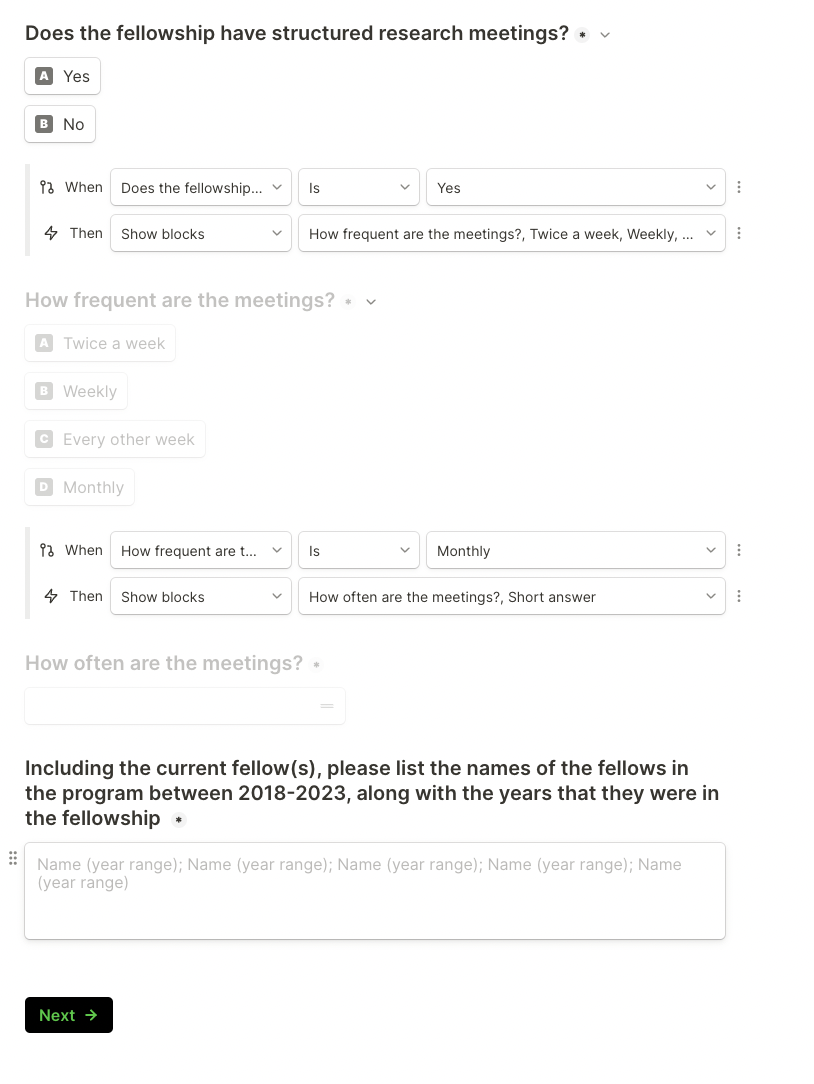

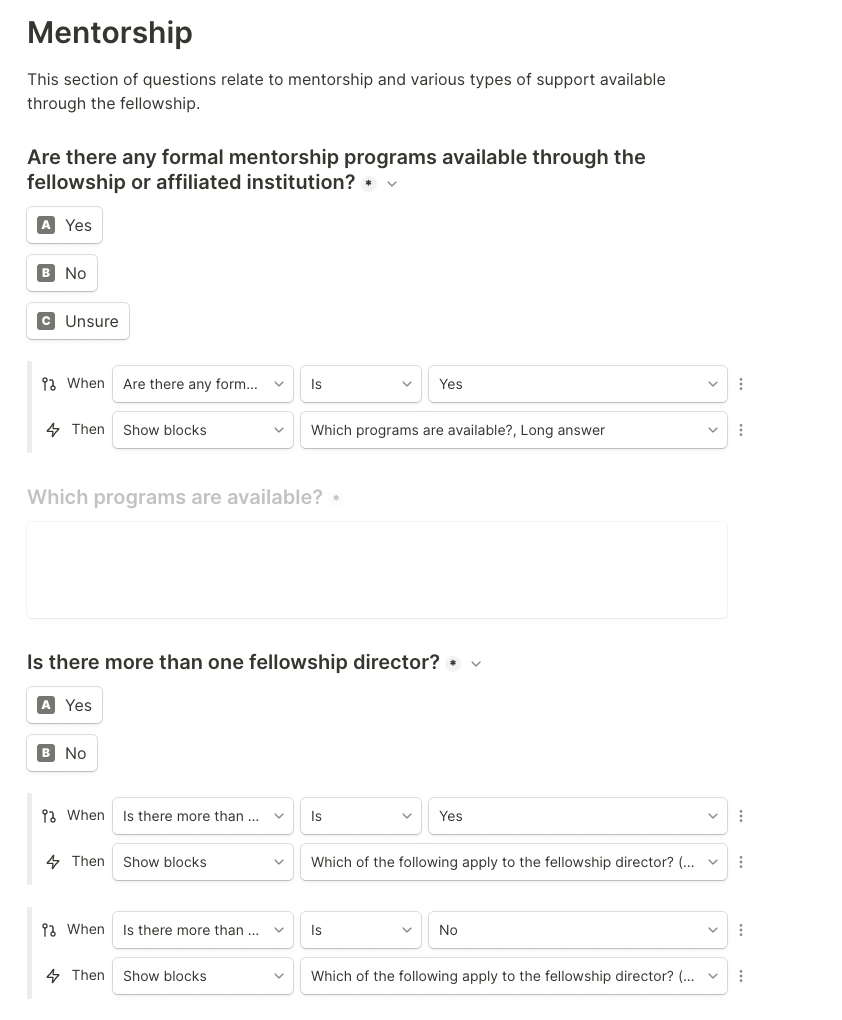

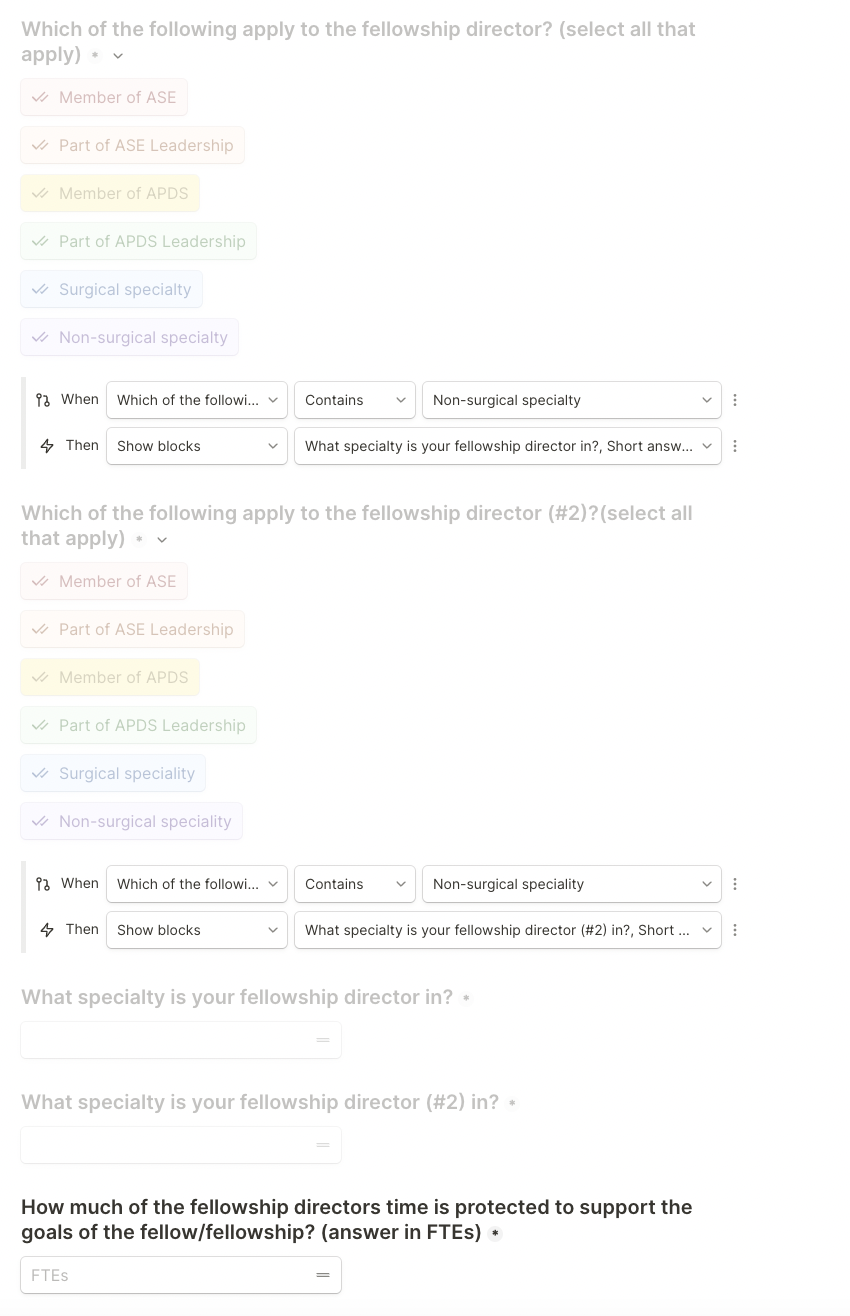

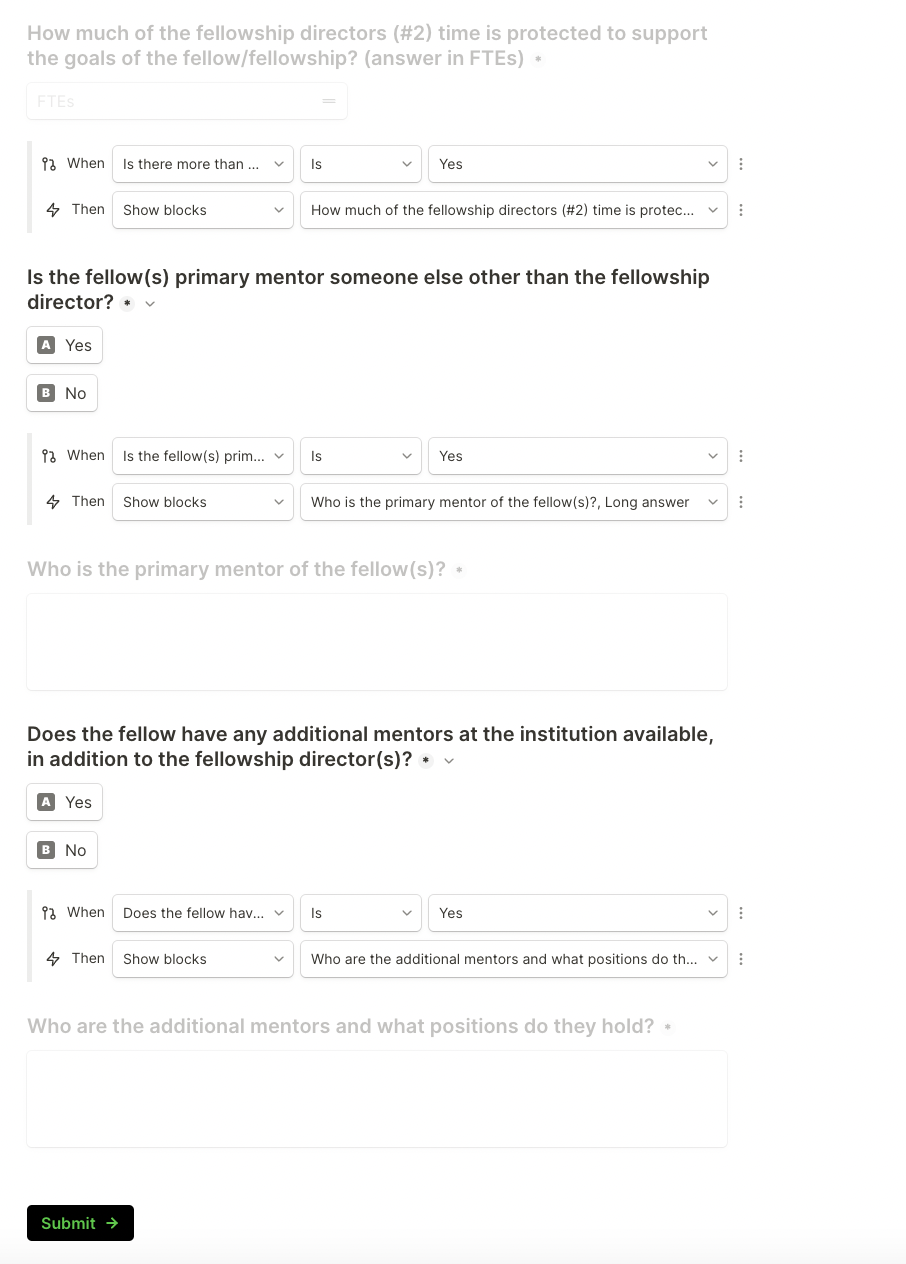
