## Supplemental B for "A Review of Surgical Education Fellowships in the United States"

***Supplementary B*:** Additional Fellowship Program Characteristics

| **Program Characteristic** | **Frequency (%)** |
| --- | --- |
| *Number of fellowship positions available per cycle (n=19)*  One position  One to two positions  Two positions  Three positions  Variable | 11 (58%)  4 (21%)  1 (5%)  1 (5%)  2 (11%) |
| *Types of positions available (n=19)*  Full-time only  Full-time and part-time  Part-time only | 15 (79%)  4 (21%)  0 (0%) |
| *Hold regular research meetings (n=19)*  Yes  No | 17 (89%)  2 (11%) |
| *Frequency of structured research meetings (n=17)*  Twice a week  Weekly  Every other week  Monthly | 2 (11.5%)  10 (59%)  2 (11.5%)  3 (18%) |
| *Fellowship directors’ national membership and leadership roles (n=24)*  ASE Member  ASE Leadership  APDS Member  APDS Leadership  ACS Member  ACS Leadership | 17 (71%)  9 (37.5%)  14 (58%)  5 (21%)  20 (83%)  4 (17%) |
